# Impact of habitual betel quid chewing on cardiovascular risk and outcomes: a systematic review

**DOI:** 10.1101/2023.11.14.23298500

**Authors:** Rodney Itaki, Shalon Taufa

**Affiliations:** American Samoa Department of Health, American Samoa; Division of Basic Medical Sciences, School of Medicine & Health Sciences, University of Papua New Guinea

**Keywords:** betel nut, betel quid, areca catechu, areca nut, cardiovascular risk, adverse cardiovascular outcome

## Abstract

**Objectives:** Habitual betel quid chewing is a leading cause of oral cancer in Asia-Pacific countries where this practice is prevalent. While health policies have focused on countering betel quid chewing concerning cancer, current policies and health promotion strategies overlook the emerging link to adverse cardiovascular outcomes. This oversight could be due to inadequate studies demonstrating the association between betel quid chewing and cardiovascular risk. To address this gap, we conducted a systematic literature review and narrative synthesis of peer-reviewed published studies showing habitual betel quid use as a cardiovascular risk factor.

**Methods:** We searched PubMed for studies assessing betel quid chewing and its impact on cardiovascular health. We included primary research on human subjects. Next, we extracted data from eligible studies and stratified by geographical location, study designs and cardiovascular outcomes. Finally, we did a narrative synthesis of the data to identify adverse cardiovascular outcomes associated with chronic betel quid use. We did not do a meta-analysis because of the different study designs, cardiovascular outcomes, and statistical measures.

**Results:** We reviewed data from 19 studies that met the inclusion criteria. Habitual betel quid chewing is associated with ischemic heart disease, obstructive coronary artery disease, acute coronary syndrome, and re-hospitalisation following an acute coronary event. Additionally, betel quid use is a risk factor for atrial fibrillation and premature ventricular contractions. Long-term betel quid consumption was associated with elevated risks of all-cause mortality, cancer-related mortality, cardiovascular diseases, and cerebrovascular diseases. Moreover, habitual betel quid users had a higher overall cardiovascular risk. The regular use of betel quid was positively correlated with arterial wall stiffness and was independently associated with heart disease in women. Habitual betel quid use is associated with hypertension.

**Conclusions:** Habitual betel quid chewing is an important cardiovascular risk factor in populations where the practice is prevalent.

## Introduction

Betel quid (Areca catechu) chewing is a common practice in the Asia-Pacific Region, though there are variations in chewing techniques across countries (1–3). This habit is known for its addictive nature, attributed to the mild euphoric effect and heightened mental alertness it bestows (4). The association between betel quid chewing and the development of oral pre-malignant and malignant lesions has been established, with the World Health Organization (WHO) classifying betel quid as a carcinogen (1). Additionally, betel quid chewing is linked to the exacerbation of asthma (5), adverse pregnancy outcomes (6), disruptions in Vitamin D metabolism (7), manganese toxicity (8), and abnormal psychological profiles (4).

Betel nut contains many compounds, with the four primary active ingredients being alkaloids: arecoline, arecaidine, guvacoline, and guvacine (4). The chemical structure of arecoline is similar to acetylcholine and stimulates muscarinic and nicotinic receptors (4). It is the primary alkaloid producing pharmacological effects such as euphoria, central nervous system stimulation, vertigo, excessive salivation, miosis, and tremor (4). Betel quid chewing also causes transient tachycardia, but the clinical implications are unknown (9). Laboratory studies suggest arecoline may be metabolised by acetylcholinesterase and carboxylesterase, the same enzymes that break down acetylcholine (10).

Recently, attention has been directed towards betel quid chewing as a metabolic and cardiovascular risk factor (11). For instance, betel quid chewing is associated with an increased risk for obesity, diabetes mellitus, metabolic syndrome, hypertension, ischemic heart disease, and arrhythmia (11). While public health interventions have primarily focused on countering betel quid chewing concerning cancer (12), current policies and health promotion strategies overlook the emerging link to adverse cardiovascular outcomes. This oversight could be due to inadequate epidemiological, clinical, and laboratory-based studies demonstrating the correlation between betel quid chewing and its impact on cardiovascular health. To address this gap, we conducted a systematic literature review and narrative synthesis of published studies that show the epidemiological link between betel quid chewing and adverse cardiovascular outcomes. The findings from our systemic review will be valuable for researchers in public health policy, health promotion and clinicians.

## Methods

### Source data

We collected data from studies indexed in PubMed. We did not search other databases (e.g. Scopus and Web of Science) because we do not have institutional subscription access. Scopus and Web of Science do not offer individual subscriptions. We did not use Google Scholar because it can produce inaccurate results (13). PubMed remains the database of choice for clinicians and researchers in biomedical sciences conducting systematic literature reviews (13).

### Search strategy and study selection

We searched PubMed using the following search terms in the title and abstracts for primary studies published between 1951 and 2021:

1. (“betel nut” OR “betel quid” OR “areca nut”)
2. (“cardiovascular disease*”)
3. (hypertension OR “high blood pressure”)
4. (“coronary artery disease*”)
5. (“ischemic heart disease*”)
6. (“ischaemic heart disease*”)
7. (atherosclerosis)
8. (arrhythmia*)
9. #1 AND #2
10. #1 AND #3
11. #1 AND #4
12. #1 AND #5
13. #1 AND #6
14. #1 AND #7
15. #1 AND #8

We accepted the results of step 9 to step 15 for screening. This resulted in 162 studies. Filtering using the PubMed automation tool produced 129 studies. We used the Preferred Reporting Items for Systemic Reviews and Meta-Analysis (PRISMA) guidelines to screen the abstracts of the final 129 studies (Figure 1.0) using pre-defined inclusion/exclusion criteria to select eligible studies (14).

**Figure 1.0.**
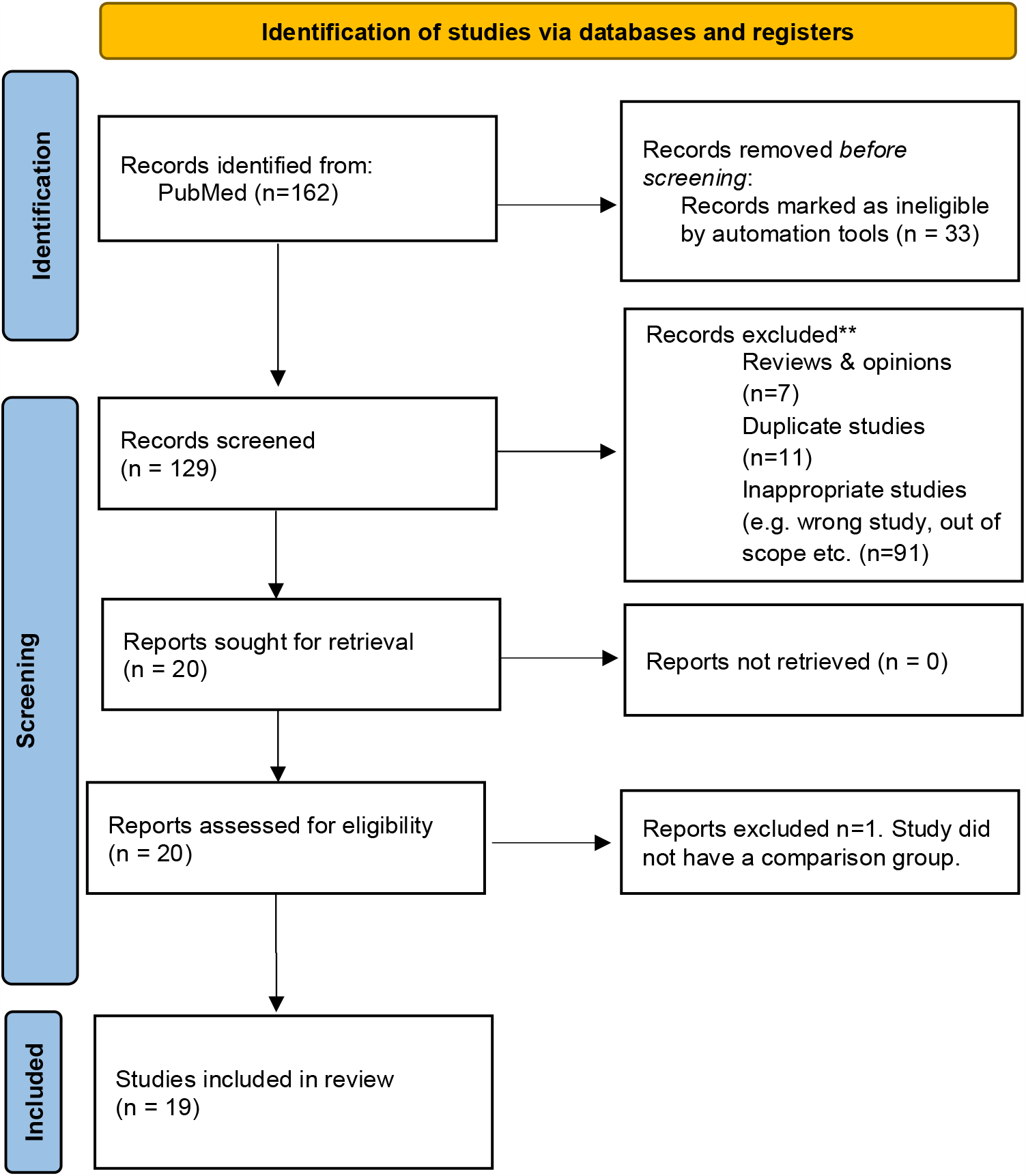
PRISMA flow diagram.

The inclusion/exclusion criteria were as follows:

#### Inclusion criteria

1. Human studies
2. Cohort (retrospective and prospective), case-control, ecological studies and cross-sectional studies
3. Peer-reviewed primary clinical or epidemiological research.

#### Exclusion criteria

1. Editorials
2. Letters, opinions and commentaries
3. Reviews (systemic or scoping)
4. Laboratory-based studies using human tissue or cells
5. Duplicate studies
6. Wrong studies or studies out of scope

We obtained the full text of all eligible studies for in-depth reading, data extraction and analysis. We excluded studies for which we were unable to retrieve full-text articles.

### Data extraction, analysis and quality assessment

The data extraction method was designed by RI using the PICO framework. All relevant data were extracted by reading full-text articles of selected studies and entered into an Excel sheet. We extracted the data independently and then combined the results for analysis. We resolved any disagreement in the data by consensus.

We assessed the quality of studies using the Critical Appraisal Skills Programme (CASP) checklist (15).

### Analytical framework

We stratified the studies by country. Next, we categorised the studies in each country by study design and extracted data relevant to cardiovascular outcomes. Finally, we compared and contrasted the cardiovascular outcomes using the PICO framework by country, study designs, and cardiovascular outcomes of each study to produce a narrative synthesis of the data. We opted not to do a meta-analysis because of the different study designs and cardiovascular outcomes.

## Results

### Study setting and study design

We extracted data from 19 studies (Figure 1). Thirteen studies were done in Taiwan (16–28), three studies done in Pakistan (29–31) and three studies in Bangladesh (32–34). There were nine cross-sectional studies(16–18,20,24–27,31), seven prospective cohort studies (19,22,28,29,32–34), two case-control studies (23,30) and one experimental design (21). Of the nine cross-sectional studies, eight were done in Taiwan (16–18,20,24–27) and one in Pakistan (31). Of the seven prospective studies, three were done in Bangladesh (32–34), three in Taiwan (19,22,28)v and one in Pakistan (29). One case-control study was done in Pakistan (30), and one was done in Taiwan (23). The experimental study was done in Taiwan (21).

#### Betel quid use and cardiovascular outcome

##### Hypertension

Three studies examined blood pressure (BP) as a cardiovascular outcome. Two were cross-sectional studies (16,25) and one prospective cohort study (32). The two cross-sectional studies were done in Taiwan (16,25), whereas the prospective cohort study was done in Bangladesh (32). Heck JE et al (2012) and Tseng (2008) showed betel quid chewing was associated with hypertension, but Chung et al (2007) showed the opposite. The risk of hypertension with betel quid chewing was independent of smoking (25). After controlling for confounding variables, betel quid users had higher odds of general hypertension (OR=1.48, 95% CI 1.04-2.10) and systolic hypertension (OR=1.55, 95% CI 1.01-2.37) (32). Notably, ex-chewers continued to have elevated BP compared to non-chewers many years after stopping the habit (32). The tendency to have low BP with betel quid chewing may be linked to genetics (16). Still, more studies are needed to provide information on the underlying pathophysiology. The persistence of hypertension among ex-betel quid chewers could be due to atherosclerosis (33) and arterial wall stiffness (27). Systemic inflammation may also play a role in persistent high blood pressure in betel quid chewers ((20).

##### Atherosclerosis

Two studies examined the impact of betel quid use on blood vessels. The first study by McClintock et al (2014) assessed carotid intima-media thickness (IMT) as an indicator of atherosclerosis and found a positive correlation between the duration and cumulative exposure of betel quid use and IMT. This association was more pronounced in men, suggesting that prolonged or high cumulative betel consumption is linked to subclinical atherosclerosis, as indicated by carotid IMT. In contrast, Wei et al (2017) investigated arterial wall stiffness and found that both ex-chewers and current chewers had an increased risk of arterial stiffness compared to non-chewers. The risk was significantly higher among individuals who had been chewing betel quid for 10 or more years and those who consumed larger daily quantities. Cumulative exposure to betel use, measured in piece-years, was positively associated with increased arterial stiffness. The results from the two studies suggest that betel quid use affects blood vessel walls. However, the underlying pathophysiological mechanisms are unknown.

##### Inflammation

Betel quid use is associated with systemic inflammation (20,31). Betel quid chewers had a higher likelihood (OR=3.23, 95% CI 2.08-5.02, p<0.001) of elevated C reactive protein (CRP) levels than non-chewers (31). Furthermore, the evidence indicates a dose-response pattern, suggesting that the quantity of betel nuts consumed may influence the magnitude of the inflammatory response (20). Additionally, individuals who chewed betel quid with tobacco were two times more likely to have elevated CRP levels. Importantly, these associations remained significant even after adjusting for potential confounding factors like age, body mass index (BMI), and education. Overall, these findings underscore the need for laboratory-based studies to elucidate the role of betel nut-induced inflammation and its role in the development of cardiovascular diseases.

##### Ischemic heart disease

Four studies investigated the relationship between betel quid and ischemic heart disease. Two studies were conducted in Pakistan (29,30) and two in Taiwan(23,26). The results from all four studies indicate betel quid consumption is a risk factor for ischemic heart disease (IHD). Regular betel quid users were 3.5 times more likely to have obstructive coronary artery disease (CAD) compared to non-users (23). Additionally, even after adjusting for other risk factors such as diabetes and hypertension, betel quid users were seven times more likely to have CAD ((30). In those who were hospitalised following an acute coronary syndrome (ACS), the risk of re-hospitalisation in one month was twice that of non-betel quid users (aHR=2.05, 95% CI 1.29-3.27) and had higher 30-day mortality (aHR=2.72, 95% CI 1.73-4.26) (29). Furthermore, older individuals are at a higher risk of subclinical IHD compared to younger consumers of betel quid (26). These findings underscore the importance of recognising betel quid use as a preventable risk factor for IHD.

##### Cerebral blood flow

We identified one experimental study assessing the effect of betel quid consumption on cerebral blood flow (21). Cerebral blood flow, blood pressure and heart rate were recorded in healthy male volunteers after chewing betel quid. Chronic chewers experienced delayed facial flushing and shorter duration, suggesting possible tolerance. The systolic BP rose, but the diastolic BP dropped two minutes after betel quid chewing in novice and chronic chewers. However, there was no statistical difference between the two groups. Heart rate rose by an average of 120% from baseline in novice and occasional users two minutes after chewing betel quid. Blood flow velocity in the external and common carotid arteries increased after betel quid chewing, while no changes occurred in the internal carotid and middle cerebral arteries. Intracranial cerebral hemodynamics remained unaffected by betel chewing. The clinical implications of these results remain unclear.

##### Arrhythmia

Betel quid use is a risk factor for arrhythmias (24) and premature ventricular contractions (PVCs) (18). In a nationwide ecological study, Tsai et al (2013) found a positive correlation (r=0.558, p=0.07) between areca quid chewing and atrial fibrillation (AF). Additionally, the odds of having AF were higher among betel quid users (OR=1.02,95% CI =1.00-1.04). On the other hand, Huang et al (2020) showed that betel quid users had a higher prevalence and burden of PVCs and a higher risk of heart failure compared to non-chewers (aOR=3.4, 95% CI 1.4-8.7). The risk of PVC was more pronounced in patients with pre-existing congestive heart failure.

##### Heart disease in women

One study evaluated gender differences in betel quid use and association with heart diseases (17). The results showed that betel quid use was more prevalent among men compared to women (31% vs 2.4%, p<0.001) (17). When comparing gender differences, the prevalence of heart disease didn’t significantly differ between men (1.5%) and women (3.2%) betel quid users (17). Additionally, betel quid use was positively associated with a higher Framingham risk score even after removing confounding factors. Interestingly, betel quid use was independently and positively associated with heart disease in women when assessed as a continuous variable but not when analysed as a categorical variable (17). Although this study suggests an association between betel quid use and heart disease in women, the results are inconclusive and warrant further investigation.

##### Cause-specific and cardiovascular mortality

Three studies investigated the impact of betel quid use on mortality (19,22,34). The first, conducted in Bangladesh, showed betel quid use was associated with a 26% higher risk of all-cause mortality and a 55% higher risk of cancer-related mortality, with no significant link to cardiovascular disease mortality (34). The second study, which had a 9.5-year follow-up period, showed betel quid chewers had an increased risk of mortality from cardiovascular conditions, cerebrovascular disease, and all causes combined, particularly with longer-term use (19). The third study demonstrated an elevated risk of CVD and overall mortality, with the risk being higher among frequent users, regardless of smoking habits (22). While all studies consistently show betel quid use with higher mortality risk, there were differences in specific outcomes and study populations.

##### Overall cardiovascular risk

Betel quid chewing contributes to cardiovascular risk (20,28). Yen et al (2008) found that betel quid chewing is associated with a 23% higher risk of CVD in Taiwanese men, even after adjusting for age and education. The study established a dose-response relationship between betel quid chewing and CVD risk, with longer duration, higher daily usage rates, and cumulative exposure intensifying the risk. Additionally, betel quid use is associated with multiple CVD risk factors, including hypertension and systemic inflammation (20).

## Discussion

Results from this systematic review suggest habitual betel quid chewing is a risk factor for poor cardiovascular outcomes. Our review primarily focused on evaluating human studies. All the studies included in our review originated from three Asian countries: Taiwan, Bangladesh, and Pakistan. The epidemiological studies demonstrate a strong association between betel quid chewing and several cardiovascular outcomes, including hypertension, ischemic events, arrhythmias, increased mortality, and a higher risk of cardiovascular diseases. However, insufficient evidence exists to determine whether the habit is an independent cardiovascular risk factor or betel quid chewing exacerbates pre-existing cardiovascular conditions.

Current epidemiological evidence suggests betel quid use is associated with hypertension. Our review contributes to the existing evidence (11). However, there is insufficient evidence to determine if this association is independent of other cardiovascular risk factors, such as smoking status. Genetic factors may also have a role (32). Additionally, atherosclerosis, arterial wall stiffness, and systemic inflammation may contribute to the persistence of high blood pressure among ex-betel quid chewers. Chronic use of betel quid and Piper betle can cause persistent sympathetic drive, contributing to hypertension (35). The relationship between betel quid chewing and blood pressure appears multifactorial and requires continued investigation.

The evidence in this review indicates betel quid is a risk factor for IHD. The risk of an ischemic event is higher in people with pre-existing CAD. Arecoline may cause coronary artery spasms due to parasympathetic effects on abnormal endothelium (36). Additionally, coronary artery ischemia may be precipitated by arecoline, similar to the action of acetylcholine (37). Long-term betel quid use also increases CAD in Taiwanese men as an independent risk factor (23). Acetylcholine causes vasodilatation in coronary arteries with normal endothelium but paradoxically induces vasoconstriction in atherosclerosed coronary arteries (37). The alkaloids in betel nut may act via a similar mechanism in normal coronary arteries but paradoxically cause vasoconstriction in atheroslerosed coronary arteries. The paradoxical coronary artery vasospasm may be the underlying pathological mechanism explaining myocardial ischemia after chewing betel nut in high-risk individuals (38).

Betel quid consumption increased blood flow velocity in the external and common carotid arteries. However, no significant changes were observed in the internal carotid and middle cerebral arteries, implying regional variations in blood flow response (21). These findings are supported by other studies showing transient tachycardia and variable blood pressure response (9,35). Crucially, intracranial cerebral hemodynamics appeared unaffected by betel chewing (21). The alkaloids, once absorbed, may be rapidly metabolised, hence the transient nature of the acute effects.

Betel quid users with pre-existing IHD have an increased risk of arrhythmias. In healthy volunteers, betel quid consumption results in a brief but rapid tachycardia lasting approximately seven minutes (9). However, chewing betel quid may induce myocardial ischemia in individuals with pre-existing IHD, leading to the onset of arrhythmias perhaps precipitated by acute transient tachycardia or coronary artery spasms (35,38). This mechanism could underlie cases of sudden death among betel quid consumers (38).

Betel quid use is also a key risk factor for metabolic syndrome (11). In patients with pre-existing conditions such as type 2 diabetes mellitus, ischemic heart disease or hypertension, habitual use of betel quid increases the risk of death from CVDs (11). Our review contributes to this body of evidence. Although the underlying pathophysiological mechanisms are unknown, computational modelling suggests arecoline interferes with lipid metabolism and possibly plays a role in the pathophysiology of atherosclerosis, hyperglycaemia, hyperlipidemia and insulin resistance (39). Betel quid-induced systemic inflammation may also be a contributing factor. Detailed laboratory-based studies are needed to explain the possible underlying pathophysiological mechanisms.

Betel quid use is consistently associated with higher mortality and overall cardiovascular risks. The specific risks varied across studies, with some showing increased risks of cancer-related mortality, cardiovascular disease mortality, CVD mortality, and overall mortality (11). Long-term use of betel products and its link to cancer development is well established. However, this review highlights the cardiovascular health consequences of chronic betel quid consumption. The impact appears to be more pronounced in individuals with pre-existing IHD or CAD disease.

There are several limitations to our review. Firstly, we extracted studies from only one database. We are uncertain whether the results would have differed if we had utilised two or three databases. We may have overlooked some studies deposited in other databases. We chose not to use Google Scholar because it is not a reliable source for systematic reviews (13). Secondly, all the studies analysed in this review are from three Asian countries, with the majority originating from Taiwan. Consequently, the results represent data from Asia, possibly limiting their generalizability.

Betel quid is also consumed in some Pacific Island countries and India, but our review did not encompass studies from these regions (3,6,40,41). Thirdly, almost all the cohort studies were conducted in Taiwan, potentially skewed towards Taiwan. Fourthly, recall bias may have influenced the results of all the studies included in our review because the authors used questionnaires to obtain data about betel nut use. Fifth, all the studies included in this review did not stratify the outcomes by socioeconomic status. We may have noticed a difference in risk between individuals from lower socioeconomic status and those from middle or high socioeconomic status. However, all the studies in this review consistently show betel quid chewing is a risk factor for CVDs. Finally, we opted not to do a meta-analysis because of the different study designs and cardiovascular outcomes. We do not think a meta-analysis would have affected our conclusions. A meta-analysis would have shown differences in risk magnitude and variability in effect sizes across the studies we reviewed but would not have changed our overall conclusions (42).

This systemic review highlighted the association between habitual betel quid chewing and CVDs. Firstly, there is an increased risk of ischemic events. Secondly, the relationship between betel quid use and blood pressure is complex and potentially influenced by genetic factors. Third, betel quid consumption adversely affects blood vessels, increasing the risk of conditions like atherosclerosis. Fourth, acute effects include temporary changes in blood flow and heart rate, increasing the risk for arrhythmias in individuals with pre-existing IHD and CAD. Fifth, gender-specific analysis showed higher male smoking rates and higher female hypertension prevalence among betel quid users, with an independent link to heart diseases in women. Finally, betel quid use was associated with increased all-cause mortality risk in long-term users. In conclusion, this review highlights that habitual betel quid use is associated with adverse cardiovascular health outcomes in regions where this habit is common.

## Supporting information

List of articles and PICO framework

## Data Availability

1. List of articles included in the review.

2. PICO framework used for data extraction and analysis.

## References

1. International Agency for Research on Cancer. IARC monographs on the evaluation of carcinogenic risks to humans, volume 85, Betel-quid and areca-nut chewing and some areca-nut-derived nitrosamines. Vol. 85. Lyon: World Health Organization; 2003.

2. Murphy KL, Herzog TA. Sociocultural Factors that Affect Chewing Behaviors among Betel Nut Chewers and Ex-Chewers on Guam. Hawaii J Med Public Health. 2015;74(12):406–11.

3. Paulino YC, Ettienne R, Novotny R, Wilkens LR, Shomour M, Sigrah C, et al. Areca (betel) nut chewing practices of adults and health behaviors of their children in the Freely Associated States, Micronesia: Findings from the Children’s Healthy Living (CHL) Program. Cancer Epidemiol. 2017 Oct;50(Pt B):234–40.

4. Osborne PG, Chou TS, Shen TW. Characterization of the Psychological, Physiological and EEG Profile of Acute Betel Quid Intoxication in Naïve Subjects. PLoS ONE. 2011 Aug 31;6(8):e23874.

5. Wang TN, Huang MS, Lin MC, Duh TH, Lee CH, Wang CC, et al. Betel Chewing and Arecoline Affects Eotaxin-1, Asthma and Lung Function. PLoS ONE. 2014 Mar 21;9(3):e91889.

6. Senn M, Baiwog F, Winmai J, Mueller I, Rogerson S, Senn N. Betel nut chewing during pregnancy, Madang province, Papua New Guinea. Drug and Alcohol Dependence. 2009 Nov 1;105(1):126–31.

7. Ogunkolade WB, Boucher BJ, Bustin SA, Burrin JM, Noonan K, Mannan N, et al. Vitamin D Metabolism in Peripheral Blood Mononuclear Cells Is Influenced by Chewing “Betel Nut” (Areca catechu) and Vitamin D Status. The Journal of Clinical Endocrinology & Metabolism. 2006 Jul 1;91(7):2612–7.

8. Al-Rmalli SW, Jenkins RO, Haris PI. Betel quid chewing as a source of manganese exposure: total daily intake of manganese in a Bangladeshi population. BMC Public Health. 2011 Dec;11(1):85.

9. Itaki R, Kevau IH, Miam B, Giva M, Mari R, Waigiebu J. Comparision of the heart rates and blood pressure of normotensive and hypertensive betel nut (areca catechu) chewers. Pac J Med Sci. 2015;14(2):16–23.

10. Giri S, Idle JR, Chen C, Zabriskie TM, Krausz KW, Gonzalez FJ. A Metabolomic Approach to the Metabolism of the Areca Nut Alkaloids Arecoline and Arecaidine in the Mouse. Chem Res Toxicol. 2006 Jun 1;19(6):818–27.

11. Yamada T, Hara K, Kadowaki T. Chewing Betel Quid and the Risk of Metabolic Disease, Cardiovascular Disease, and All-Cause Mortality: A Meta-Analysis. PLoS ONE. 2013 Aug 5;8(8):e70679.

12. Mehrtash H, Duncan K, Parascandola M, David A, Gritz ER, Gupta PC, et al. Defining a global research and policy agenda for betel quid and areca nut. The Lancet Oncology. 2017 Dec;18(12):e767–75.

13. Falagas ME, Pitsouni EI, Malietzis GA, Pappas G. Comparison of PubMed, Scopus, Web of Science, and Google Scholar: strengths and weaknesses. FASEB j. 2008 Feb;22(2):338–42.

14. Page MJ, McKenzie JE, Bossuyt PM, Boutron I, Hoffmann TC, Mulrow CD, et al. The PRISMA 2020 statement: an updated guideline for reporting systematic reviews. BMJ. 2021 Mar 29;n71.

15. Critical Appraisal Skills Programme [Internet]. 2023 [cited 2023 Sep 22]. Critical Appraisal Skills Programme. Available from: https://casp-uk.net/casp-tools-checklists/

16. Chung FM, Shieh TY, Yang YH, Chang DM, Shin SJ, Tsai JCR, et al. The role of angiotensin-converting enzyme gene insertion/deletion polymorphism for blood pressure regulation in areca nut chewers. Translational Research. 2007 Jul;150(1):58–65.

17. Guh JY, Chen HC, Tsai JF, Chuang LY. Betel-quid use is associated with heart disease in women. Am J Clin Nutr. 2007;85):1229–35.

18. Huang TC, Wu WT, Chen YC, Yang FM, Tsai WC, Lee CH. Betel-Quid Chewing, Heart Failure, and Premature Ventricular Contractions in Patients with Cardiopulmonary Symptoms. Int J Environ Res Public Health. 2020 Oct 14;17(20):7472.

19. Lan TY, Chang WC, Tsai YJ, Chuang YL, Lin HS, Tai TY. Areca Nut Chewing and Mortality in an Elderly Cohort Study. Am J Epidemiol. 2007 Jan 12;165(6):677–83.

20. Lin SH, Liao YS, Huang SH, Liao WH. Relationship between betel quid chewing and risks of cardiovascular disease in older adults: A cross-sectional study in Taiwan. Drug and Alcohol Depend. 2014 Aug;141:132–7.

21. Lin SK, Chang YJ, Ryu SJ, Chu NS. Cerebral Hemodynamic Responses to Betel Chewing: A Doppler Study: Clin Neuropharmacol. 2002 Sep;25(5):244–50.

22. Lin WY, Chiu TY, Lee LT, Lin CC, Huang CY, Huang KC. Betel nut chewing is associated with increased risk of cardiovascular disease and all-cause mortality in Taiwanese men. Am J Clin Nutr. 2008 May;87(5):1204–11.

23. Tsai WC, Wu MT, Wang GJ, Lee KT, Lee CH, Lu YH, et al. Chewing areca nut increases the risk of coronary artery disease in Taiwanese men: a case-control study. BMC Public Health. 2012 Dec;12(1):162.

24. Tsai WC, Chen CY, Kuo HF, Wu MT, Tang WH, Chu CS, et al. Areca Nut Chewing and Risk of Atrial Fibrillation in Taiwanese Men: A Nationwide Ecological Study. Int J Med Sci. 2013;10(7):804–11.

25. Tseng CH. Betel Nut Chewing Is Associated with Hypertension in Taiwanese Type 2 Diabetic Patients. Hypertens Res. 2008;31(3):417–23.

26. Tseng CH. Betel Nut Chewing and Subclinical Ischemic Heart Disease in Diabetic Patients. Cardiol Res Pract. 2011;2011:1–5.

27. Wei YT, Chou YT, Yang YC, Chou CY, Lu FH, Chang CJ, et al. Betel nut chewing associated with increased risk of arterial stiffness. Drug and Alcohol Depend. 2017 Nov;180:1–6.

28. Yen AMF, Chen LS, Chiu YH, Boucher BJ, Chen THH. A prospective community-population-registry– based cohort study of the association between betel-quid chewing and cardiovascular disease in men in Taiwan (KCIS no. 19). Am J Clin Nutr. 2008 Jan;87(1):70–8.

29. Karim MT, Inam S, Ashraf T, Shah N, Adil SO, Shafique K. Areca Nut Chewing and the Risk of Rehospitalization and Mortality Among Patients With Acute Coronary Syndrome in Pakistan. J Prev Med Public Health. 2018 Mar 31;51(2):71–82.

30. Khan MS, Bawany FI, Ahmed MU, Hussain M, Khan A, Lashari MN. Betel Nut Usage Is a Major Risk Factor for Coronary Artery Disease. Glob J Health Sci. 2014;6(2):189–95.

31. Shafique K, Mirza S, Vart P, Memon A, Arain M, Tareen M, et al. Areca nut chewing and systemic inflammation: evidence of a common pathway for systemic diseases. J Inflamm. 2012;9(1):22.

32. Heck JE, Marcotte EL, Argos M, Parvez F, Ahmed A, Islam T, et al. Betel quid chewing in rural Bangladesh: prevalence, predictors and relationship to blood pressure. Int J Epidemiol. 2012 Apr 1;41(2):462–71.

33. McClintock TR, Parvez F, Wu F, Wang W, Islam T, Ahmed A, et al. Association between betel quid chewing and carotid intima-media thickness in rural Bangladesh. Int J Epidemiol. 2014 Aug;43(4):1174–82.

34. Wu F, Parvez F, Islam T, Ahmed A, Rakibuz-Zaman M, Hasan R, et al. Betel quid use and mortality in Bangladesh: a cohort study. Bull World Health Organ. 2015 Oct 1;93(10):684–92.

35. Garg A, Chaturvedi P, Gupta PC. A review of the systemic adverse effects of areca nut or betel nut. Indian J Med Paediatr Oncol. 2014 Jan;35(01):3–9.

36. Hung DZ, Deng JF. Acute myocardial infarction temporally related to betel nut chewing. Vet Hum Toxicol. 1998 Feb;40(1):25–8.

37. Ludmer PL, Selwyn AP, Shook TL, Wayne RR, Mudge GH, Alexander RW, et al. Paradoxical vasoconstriction induced by acetylcholine in atherosclerotic coronary arteries. N Engl J Med. 1986 Oct 23;315(17):1046–51.

38. Ying-Chih Chen, Hsiang-Chun Lee, Hei-Hwa Lee, Ho-Ming Su, Tsung-Hsien Lin, Po-Chao Hsu. Areca Nut Chewing Complicated with Non-Obstructive and Obstructive ST Elevation Myocardial Infarction. Acta Cardiologica Sinica. 2016 Jan 31;32(1).

39. Choudhury MD, Chetia P, Choudhury KD, Talukdar AD, Datta-choudhari M. Atherogenic effect of Arecoline: A computational study. Bioinformation. 2012 Mar 17;8(5):229–32.

40. Dalisay F, Pokhrel P, Buente W, Kawabata Y. Exposure to tobacco and betel nut content on social media, risk perceptions, and susceptibility to peer influence among early adolescents in Guam. Addict Behav Rep. 2022 Jan 3;15:100405.

41. Paulino YC, Hurwitz EL, Ogo JC, Paulino TC, Yamanaka AB, Novotny R, et al. Epidemiology of areca (betel) nut use in the Mariana Islands: Findings from the University of Guam/University of Hawaii Cancer Center Partnership Program. Cancer Epidemiol. 2017 Oct;50(Pt B):241–6.

42. Cheung MWL, Vijayakumar R. A Guide to Conducting a Meta-Analysis. Neuropsychol Rev. 2016 Jun;26(2):121–8.

